# Genetic Correlation Between Brain Imaging Phenotypes and Externalizing Behavior: A Large-Scale LDSC Analysis of UK Biobank IDPs

**DOI:** 10.64898/2026.07.12.26357891

**Authors:** Mengman Wei, Qian Peng

## Abstract

Externalizing has been associated with differences in brain structure and function; however, it remains unclear whether these associations reflect shared common-variant genetic influences.

Cross-trait linkage disequilibrium score regression was used to estimate genome-wide genetic correlations between externalizing genome-wide association study (GWAS) results and 3,935 brain imaging-derived phenotypes from the UK Biobank BIG40 resource. The imaging phenotypes covered structural magnetic resonance imaging (MRI), diffusion MRI, susceptibility-weighted imaging, resting-state functional MRI, and task-based functional MRI. Results were included in the primary analysis when the imaging phenotype had positive single-nucleotide polymorphism (SNP) heritability, a heritability *Z* statistic of at least 1.96, a mean GWAS chi-square statistic of at least 1.02, at least 200,000 regression SNPs, and a complete LDSC result without a fatal error. Technical imaging quality-control phenotypes were excluded from biological inference. Individual results were corrected using the Benjamini–Hochberg false discovery rate procedure. Aggregated Cauchy association tests (ACATs) were used to evaluate evidence across all imaging phenotypes and within predefined imaging categories. Statistical power, simultaneous confidence bounds, and alternative quality-control definitions were examined in sensitivity analyses.

Of the 3,935 imaging phenotypes, 3,716 produced estimable genetic correlations, 2,980 met the primary LDSC quality-control criteria, and 2,967 were classified as biological imaging phenotypes. No individual phenotype survived false discovery rate correction. The smallest unadjusted *P* value was 0.0005, and the minimum adjusted *q* value was 0.486. The distribution of genetic correlations was centered near zero, with a median genetic correlation of 0.0014 and a median absolute genetic correlation of 0.0338. ACAT provided no evidence of an aggregate association across all biological imaging phenotypes (*P* = 0.302), and no predefined imaging category survived multiple-testing correction. The median minimum detectable genetic correlation at 80% power was 0.216. Bonferroni-adjusted simultaneous confidence intervals were fully contained within the interval [*−*0.30, 0.30] for 80.0% of phenotypes in the primary analysis and 88.0% under the stringent heritability quality-control definition. Broad and stringent sensitivity analyses produced the same overall conclusions.

In this study, no statistically robust evidence of genome-wide genetic correlations between externalizing and individual UK Biobank brain imaging phenotypes was found. Nevertheless, small, localized, mixed-direction, or developmentally specific genetic effects remain possible.

## Introduction

Externalizing behavior describes a broad tendency toward difficulties in behavioral and emotional regulation, including antisocial behavior, aggression, attention problems, impulsivity, risk-taking, and related traits. These behaviors frequently co-occur and are partly influenced by shared genetic factors. A large multivariate genome-wide association study (GWAS) of approximately 1.5 million individuals identified a common genetic dimension underlying multiple traits related to self-regulation, substance use, and behavioral disinhibition [1, 2].

Differences in brain structure and function may represent one pathway through which genetic liability contributes to externalizing behavior. Previous neuroimaging studies have reported associations between externalizing-related traits and measures of cortical morphology, subcortical structure, white-matter organization, and functional brain networks [3, 4]. For example, the prefrontal cortex and related cortical regions have been associated with inhibitory control and other externalizing-related traits [4]. Nevertheless, an observed association between a brain measure and externalizing behavior does not necessarily indicate that the two traits share the same genetic causes. Associations between the brain and behavior may also arise from environmental exposures, developmental processes, consequences of externalizing behavior, or interactions between genetic and environmental factors.

Large imaging-genetics datasets provide an opportunity to investigate whether brain phenotypes and externalizing behavior share common genetic influences. The UK Biobank imaging-genetics resource provides GWAS summary statistics for 3,935 multimodal brain imaging-derived phenotypes [5]. These phenotypes include regional brain volumes, cortical surface area and thickness, tissue-intensity measures, white-matter microstructure, resting-state functional measures, and task-related brain activation. This broad phenotypic coverage allows genetic overlap to be examined across the brain rather than being restricted to a small number of selected regions or imaging measures.

Cross-trait linkage disequilibrium score regression (LDSC) can be used to estimate genome-wide genetic correlations between two traits using GWAS summary statistics [6]. A genetic correlation, denoted by *r*_*g*_, measures the extent to which common genetic variants influence two traits in the same or opposite directions. A positive genetic correlation indicates that genetic variants associated with higher values of one trait tend to be associated with higher values of the other trait, whereas a negative genetic correlation indicates effects in opposite directions. Cross-trait LDSC can also account for potential overlap between the samples included in the two GWASs [6].

In the present study, cross-trait LDSC was used to estimate genome-wide genetic correlations between externalizing behavior [1, 2] and all 3,935 brain imaging-derived phenotypes available from the UK Biobank resource [5]. We hypothesized that externalizing would show detectable genetic overlap with at least some brain imaging phenotypes, particularly measures of cortical morphology, subcortical structure, white-matter microstructure, and functional-network organization. Because thousands of correlated imaging phenotypes were examined, associations that survived correction for multiple comparisons were considered confirmatory, whereas associations based only on uncorrected *P* values were considered exploratory.

## Methods

### Study Design

We used publicly available genome-wide association study (GWAS) summary statistics to examine whether externalizing behavior [1, 2] and brain imaging phenotypes share common genetic influences. Genetic correlations were estimated between one externalizing GWAS and 3,935 brain imaging-derived phenotypes from the UK Biobank BIG40 resource [5].

The analysis consisted of four main stages. First, the externalizing and imaging GWAS summary statistics were converted to a common format compatible with linkage disequilibrium score regression (LDSC). Second, cross-trait LDSC was used to estimate the genetic correlation between externalizing and each imaging phenotype. Third, results that did not meet the prespecified quality-control criteria were excluded. Finally, evidence at the individual-phenotype, global, and imaging-category levels was evaluated using multiple-testing correction, omnibus tests, power calculations, and sensitivity analyses.

No individual-level genetic or imaging data were analyzed in the present study.

### Externalizing GWAS Data Source

Externalizing GWAS summary statistics were obtained from the International Externalizing Consortium. The file contained summary statistics for the externalizing factor after excluding data contributed by 23andMe. We therefore refer to this dataset as EXT-minus-23andMe [1, 2].

The original externalizing GWAS used genomic structural equation modeling to identify a shared genetic factor across seven externalizing-related traits: attention-deficit/hyperactivity disorder, problematic alcohol use, lifetime cannabis use, reverse-coded age at first sexual intercourse, number of sexual partners, general risk tolerance, and lifetime smoking initiation [1]. The original analysis included approximately 1.5 million individuals of European ancestry. The EXT-minus-23andMe dataset was generated by applying the same externalizing factor model after removing restricted 23andMe data from the contributing smoking and cannabis GWASs [2]. The reduced model showed a genetic structure and pattern of genetic correlations similar to those of the original model. The International Externalizing Consortium requires studies using this reduced release to cite both the original externalizing GWAS and the study describing the reduced summary statistics.

The effective sample size reported in the externalizing summary statistics was 1,045,957. The original file included the SNP identifier, chromosome, genomic position, effect and non-effect alleles, effect-allele frequency, effect estimate, standard error, *Z* statistic, *P* value, sample size, and an indicator of possible outlier effects.

For LDSC preparation, the following fields were used: SNP identifier, effect allele, non-effect allele, signed effect estimate, *P* value, and effective sample size. These fields were standardized as `SNP`, `A1`, `A2`, `BETA`, `P`, and `N`, respectively.

### UK Biobank Brain Imaging GWAS Data Source

Brain imaging GWAS summary statistics were obtained from the UK Biobank BIG40 resource [5]. BIG40 contains GWAS results for 3,935 imaging-derived phenotypes obtained from multimodal brain imaging.

The imaging phenotypes included measures derived from structural T1-weighted magnetic resonance imaging (MRI), cortical and subcortical segmentation, diffusion MRI, susceptibility-weighted MRI, resting-state functional MRI, and task functional MRI. Examples included regional brain volumes, cortical thickness, cortical surface area, tissue intensity, white-matter microstructure, resting-state node amplitudes, functional connectivity, and task-related activation.

The BIG40 study used a discovery sample of 22,138 participants and a replication sample of 11,086 participants. Combined sample sizes differed across phenotypes because not every participant had usable data for every imaging modality. The official BIG40 phenotype catalogue was therefore used to obtain the combined sample size, `N(all)`, separately for each phenotype. The BIG40 study and data server provide GWAS summary statistics and metadata for all 3,935 phenotypes.

The phenotype metadata contained the BIG40 phenotype number, UK Biobank field identifier, phenotype short name, full phenotype description, imaging category, measurement unit, discovery sample size, replication sample size, combined sample size, and SNP-heritability estimate.

The 3,935 records included both biological brain phenotypes and technical measures of image quality. Technical quality-control measures included head motion, image-alignment discrepancy, signal-to-noise measures, and diffusion-image outlier counts. These technical measures were retained when assessing the completeness of the analysis but were excluded from the primary biological inference.

### Preparation of the Externalizing GWAS Summary Statistics

The externalizing GWAS summary statistics were prepared using `munge sumstats.py` [7] from LDSC version 1.0.0 [8], running under Python version 2.7.11 [7, 9].

The following columns were supplied explicitly: SNP identifier, effect allele, non-effect allele, signed beta coefficient, *P* value, and sample size. These columns were standardized as `SNP`, `A1`, `A2`, `BETA`, `P`, and `N`, respectively. The beta coefficient was treated as a signed association statistic with a null value of zero.

Variants were restricted to the HapMap3 SNP list distributed with the European-ancestry LDSC reference data [7]. Restriction to HapMap3 variants is commonly used in LDSC analyses because these variants are generally well imputed and have reliable linkage disequilibrium information.

The externalizing summary statistics were processed using the following LDSC options:

~~~
--snp SNP
--a1 A1
--a2 A2
--signed-sumstats BETA,0
--p P
--N-col N
--merge-alleles w_hm3.snplist
~~~

The resulting LDSC-formatted externalizing file contained the SNP identifier, sample size, *Z* statistic, effect allele, and non-effect allele. The same externalizing file was used in every pairwise genetic-correlation analysis.

### Preparation of the BIG40 Imaging GWAS Summary Statistics

Brain imaging GWAS summary statistics were obtained from the UK Biobank BIG40 resource [5]. Each raw BIG40 file contained the following required fields: `rsid, a1, a2, beta, and pval(-log10)`.

The SNP identifier and allele columns were renamed `SNP, A1`, and `A2`. The original files reported *P* values as negative base-10 logarithms. Let *L* = *−* log_10_(*P*) denote the reported value. Ordinary *P* values were recovered as

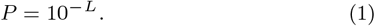

Each imaging phenotype was assigned its official combined sample size, `N(all)`, from the BIG40 phenotype catalogue. Because the source summary-statistic files did not contain SNP-specific sample sizes, the same phenotype-level sample size was assigned to every SNP within a given imaging GWAS.

Rows were removed when the SNP identifier, allele, beta coefficient, *P* value, or sample size was missing or nonfinite. Valid *P* values were required to satisfy

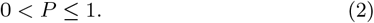

Values that were numerically equal to zero or smaller than the supported floating-point range were set to

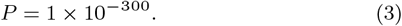

This lower bound was used only to prevent numerical errors when converting extremely small *P* values to *Z* statistics.

The imaging summary statistics were then restricted to HapMap3 variants [7]. Only the standard nucleotide alleles A, C, G, and T were retained. Variants were removed if the two alleles were identical or if they formed a strand-ambiguous pair, specifically A/T, T/A, C/G, or G/C.

The BIG40 source files included beta coefficients and *P* values but did not provide a directly usable signed *Z* statistic. Therefore, the absolute *Z* statistic was calculated from the two-sided *P* value, and its direction was determined from the sign of the beta coefficient:

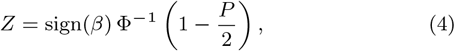

where *β* is the GWAS beta coefficient, sign(*β*) equals +1 for a positive beta and *−*1 for a negative beta, and Φ^*−*1^ is the inverse cumulative distribution function of the standard normal distribution.

When *β* = 0, *Z* was set to zero.

The final LDSC input file for each imaging phenotype contained the following columns:

`SNP N Z A1 A2`

A BIG40 summary-statistic file was required to contain at least 200,000 retained variants before it was passed to LDSC. This check was intended to identify serious harmonization problems or inadequate overlap with the HapMap3 reference set.

All BIG40 processing was completed using custom Python and AWK scripts that applied the same filtering and conversion rules to every imaging phenotype.

### Linkage Disequilibrium Reference Data

The analysis used European-ancestry LD scores from the LDSC `eur_w_ld_chr` reference dataset. These LD scores were generated using European-ancestry reference samples from the 1000 Genomes Project [10].

Analyses were restricted to the HapMap3 variants contained in `w_hm3.snplist` [7]. For regression weights, the pipeline preferentially used the HapMap3 weight files that excluded the extended major histocompatibility complex region when these files were available. Otherwise, the corresponding weights distributed with `eur_w_ld_chr` were used.

The exact LD-score and regression-weight paths used in each analysis were recorded in the LDSC log files and retained with the archived analysis outputs.

### Cross-Trait Genetic-Correlation Analysis

Cross-trait LDSC was used to estimate the genome-wide genetic correlation between externalizing [1, 2] and each BIG40 imaging phenotype [5].

For every pairwise analysis, the brain imaging phenotype was entered as phenotype 1, and externalizing was entered as phenotype 2.

Cross-trait LDSC estimates genetic covariance from the association between SNP LD scores and the products of the *Z* statistics from the two GWASs. The genetic covariance is standardized using the SNP heritabilities of the two phenotypes to obtain the genetic correlation, *r*_*g*_ [8].

For each imaging phenotype, the following results were extracted: the genetic correlation, *r*_*g*_; the standard error of *r*_*g*_; the *Z* statistic; the two-sided *P* value; SNP-heritability estimates and standard errors; univariate LDSC intercepts; and the cross-trait genetic-covariance intercept.

The null hypothesis for each individual test was

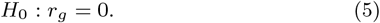

The LDSC intercepts and cross-trait intercept were estimated freely rather than fixed to one or zero. This allowed LDSC to account for possible sample overlap and correlated sources of GWAS confounding.

### Quality Control of the LDSC Results

A genetic-correlation result was first classified as structurally valid when it met all of the following conditions:

- the estimated *r*_*g*_ was finite;
- the standard error was finite and greater than zero; and
- the *P* value satisfied 0 *< P ≤* 1.

Additional quality-control criteria were then applied to the brain imaging GWAS. A phenotype was included in the primary analysis only when all of the following conditions were satisfied:

- the estimated SNP heritability was greater than zero;
- the SNP-heritability *Z* statistic was at least 1.96;
- the mean GWAS chi-square statistic was at least 1.02;
- at least 200,000 variants were retained in the LDSC regression;
- the LDSC log contained a completed genetic-correlation analysis; and
- the LDSC log contained no fatal processing error. The SNP-heritability *Z* statistic was calculated as

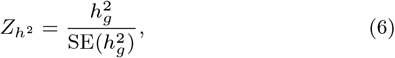

where 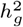 is the observed-scale SNP-heritability estimate and SE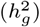 is its standard error.

The heritability and mean chi-square criteria were used to remove imaging GWASs with insufficient polygenic signal for stable genetic-correlation estimation. The regression-SNP threshold was used to identify potential problems with variant matching, harmonization, or the reference data. These were prespecified operational quality-control criteria informed by LDSC diagnostic guidance and were not treated as universal biological thresholds [11].

After LDSC quality control, phenotypes classified by BIG40 as technical imaging-quality measures were excluded from biological inference. These measures included head motion, image-alignment discrepancy, signal-to-noise measures, and diffusion-image outlier counts. The remaining imaging phenotypes formed the primary biological analysis set.

### Individual Statistical Tests and Multiple-Testing Correction

Ordinary 95% confidence intervals for individual genetic correlations were calculated as

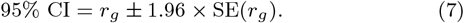

Individual *P* values were corrected across all eligible biological imaging phenotypes using the Benjamini–Hochberg false-discovery-rate (FDR) procedure [12]. A result was considered statistically significant when

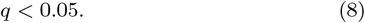

The Benjamini–Hochberg procedure was selected as the primary multiple-testing correction because it controls the expected proportion of false discoveries among the results declared significant [12].

Bonferroni correction was applied as a more conservative secondary analysis [13]. The Bonferroni-adjusted *P* value was calculated as

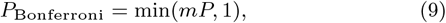

where *m* is the total number of phenotypes in the relevant analysis. For the primary analysis of 2,967 biological phenotypes, the Bonferroni significance threshold was

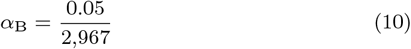

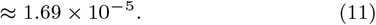

Results with an unadjusted *P <* 0.05 but an FDR-adjusted *q ≥* 0.05 were described as nominal or exploratory. They were not interpreted as statistically confirmed discoveries.

### Global and Imaging-Category Omnibus Tests

The Aggregated Cauchy Association Test (ACAT) was used to evaluate whether statistical evidence accumulated across multiple imaging phenotypes [14]. ACAT combines *P* values and is designed to remain computationally efficient when the input tests are correlated.

All phenotypes were assigned equal weights. For a group containing *m* phenotypes, the ACAT statistic was calculated as

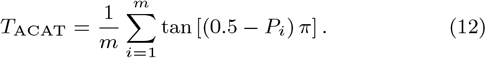

The combined *P* value was calculated as

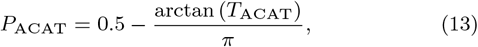

where *P*_*i*_ is the *P* value for the *i*th imaging phenotype.

For numerical stability, individual *P* values were restricted to the interval

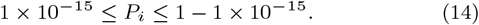

ACAT was applied at two levels:

1. across all eligible biological imaging phenotypes; and
2. separately within each of the 16 predefined biological BIG40 imaging categories.

The 16 category-level ACAT *P* values were corrected using both the Benjamini–Hochberg and Bonferroni procedures [12, 13].

To determine whether an omnibus result was driven by only a small number of phenotypes, ACAT was repeated after

- removing the phenotype with the smallest individual *P* value; and
- removing the five phenotypes with the smallest individual *P* values.

ACAT evaluates whether a group contains aggregate statistical evidence, but it does not estimate a common genetic-correlation magnitude or direction [14]. We therefore also summarized the following descriptive measures for each imaging category:

- median *r*_*g*_;
- mean *r*_*g*_;
- median absolute *r*_*g*_;
- percentage of positive *r*_*g*_ estimates;
- number of results with *P <* 0.05; and
- number of results with *P <* 0.01.

These category summaries were interpreted descriptively because imaging phenotypes within the same category were correlated.

### Power Analysis

For each imaging phenotype, we calculated the approximate minimum absolute genetic correlation detectable with 80% power at the Bonferroni-corrected significance level.

The Bonferroni-adjusted significance level was

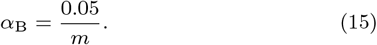

The minimum detectable effect was calculated as

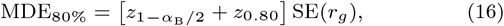

where

- *m* is the number of phenotypes in the relevant analysis;
- 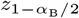 is the two-sided Bonferroni critical value;
- *z*_0.80_ is the 80th percentile of the standard normal distribution; and
- SE(*r*_*g*_) is the standard error of the estimated genetic correlation.

This calculation was based on a normal approximation and provided a phenotype-specific measure of statistical sensitivity. A larger standard error resulted in a larger minimum detectable correlation and, therefore, lower statistical power.

We summarized the number and percentage of phenotypes with approximately 80% power to detect absolute genetic correlations of

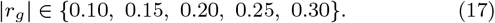

### Simultaneous Confidence Intervals

Bonferroni-adjusted simultaneous confidence intervals were calculated to determine which effect magnitudes could be excluded while controlling the family-wise error rate across all phenotypes [13].

The simultaneous confidence interval for each phenotype was

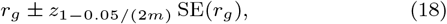

where *m* is the number of phenotypes in the relevant analysis.

For each phenotype, we assessed whether the complete simultaneous confidence interval was contained within each of the following ranges:

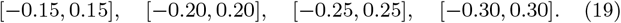

For example, if the complete simultaneous confidence interval was contained within [*−*0.30, 0.30], genetic correlations greater than 0.30 in absolute magnitude were considered inconsistent with the observed estimate under the simultaneous-confidence procedure.

These analyses were used to distinguish between two types of nonsignificant results:

- an imprecise result that could not rule out a moderate or large correlation; and
- a sufficiently precise result that provided evidence against a moderate or large correlation.

### Sensitivity Analyses

Three phenotype-inclusion definitions were evaluated.

### Primary Analysis

The primary analysis included biological imaging phenotypes that met all prespecified LDSC quality-control criteria:

- positive SNP heritability;
- heritability *Z* statistic of at least 1.96;
- mean chi-square statistic of at least 1.02;
- at least 200,000 regression SNPs;
- complete LDSC output; and
- no fatal processing error.

### Broad Sensitivity Analysis

The broad analysis included all biological imaging phenotypes with

- a finite *r*_*g*_;
- a finite and positive standard error; and
- a valid *P* value.

No minimum heritability *Z* statistic or mean chi-square threshold was required in this analysis.

### Stringent Sensitivity Analysis

The stringent analysis included biological imaging phenotypes with

- an SNP-heritability *Z* statistic of at least 3;
- a mean chi-square statistic of at least 1.02; and
- at least 200,000 LDSC regression SNPs.

For each analysis definition, we recalculated

- individual Benjamini–Hochberg *q* values;
- Bonferroni-adjusted *P* values;
- the global ACAT *P* value;
- category-level ACAT *P* and *q* values;
- median and median absolute genetic correlations;
- minimum detectable effects; and
- simultaneous confidence intervals.

The sensitivity analyses were used to determine whether the main findings depended on the specific SNP-heritability quality-control threshold.

### Software and Reproducibility

Externalizing summary-statistic munging and cross-trait genetic-correlation analyses were performed using LDSC version 1.0.0 under Python version 2.7.11 [8, 9]. The BIG40 summary statistics were processed using custom Python 2.7.11 and GNU AWK scripts that performed HapMap3 filtering, allele checks, *P* -value conversion, and signed-*Z* calculation.

Post-analysis data processing and statistical analyses were conducted using Python version 3.8.3 [15], with

- pandas version 2.0.3 for tabular data management [16];
- NumPy version 1.22.4 for array operations and numerical calculations [17];
- SciPy version 1.6.3 for standard-normal quantiles and related statistical functions [18]; and
- Matplotlib version 3.7.1 for figure generation [19].

The Benjamini–Hochberg, Bonferroni, ACAT, power, and simultaneous-confidence-interval calculations were implemented in custom Python scripts according to the procedures described above.

All 3,935 expected BIG40 phenotypes were recorded in a manifest before analysis. The pipeline checked that

- all required input files were present;
- gzip-compressed files passed integrity checks;
- each LDSC log reported successful completion;
- each completed log contained a genetic-correlation result;
- each extracted result table contained the required columns; and
- the expected number of result files was produced.

The final analysis directories were frozen on July 11, 2026. SHA-256 checksums were generated for all archived files to document the exact inputs and outputs used in the manuscript.

## Results

### Analysis Completion and Quality Control

Genetic-correlation analyses were attempted for all 3,935 BIG40 phenotypes. All expected result files were produced, and 3,716 phenotypes yielded structurally valid genetic-correlation estimates.

Among the structurally valid results, 736 did not meet the primary LDSC quality-control criteria. Of these, 154 failed only the brain SNP-heritability *Z*-statistic threshold, 97 failed only the mean chi-square threshold, and 485 failed both criteria. No phenotype failed because fewer than 200,000 regression SNPs were retained, and none of the included analyses had an incomplete or fatally terminated LDSC log.

A total of 2,980 phenotypes passed the primary LDSC quality-control criteria. Thirteen technical imaging quality-control phenotypes were subsequently excluded, leaving 2,967 biological imaging-derived phenotypes in the primary analysis.

### Individual Genetic Correlations

No individual brain imaging phenotype showed a statistically significant genetic correlation with externalizing after false-discovery-rate (FDR) or Bonferroni correction. The smallest unadjusted *P* value was 0.0005, whereas the minimum FDR-adjusted *q* value was 0.486.

The distribution of genetic-correlation estimates was centered near zero, with a median of

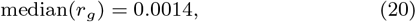

and a median absolute genetic correlation of

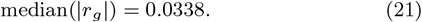

The median standard error was 0.0419.

There were 204 phenotypes with nominal *P <* 0.05, 49 with *P <* 0.01, and four with *P <* 0.001. These results were treated as exploratory because none survived correction for multiple comparisons.

The two highest-ranked exploratory results were as follows:

1. **Left medial-dorsal lateral thalamic nucleus volume:** *r*_*g*_ = 0.1155, SE = 0.0331, 95% CI = [0.0506, 0.1804], with *P* = 0.0005 and *q* = 0.486.
2. **Resting-state fMRI ICA100 node 19 amplitude:**

*r*_*g*_ = *−*0.1779, SE = 0.0508, 95% CI = [*−*0.2775, *−*0.0783], with *P* = 0.0005 and *q* = 0.486.

Other top-ranked exploratory phenotypes included right cerebral-peduncle orientation dispersion, right gyrus rectus thickness, right putamen T2*, cerebral-peduncle diffusivity, inferior frontal cortical measures, and several resting-state connectivity phenotypes. None represented a statistically significant discovery after correction for multiple comparisons.

### Global and Category-Level Evidence

There was no aggregate evidence across all 2,967 biological imaging phenotypes:

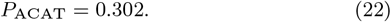

The global ACAT *P* value increased to 0.352 after removal of the phenotype with the smallest individual *P* value and to 0.529 after removal of the five phenotypes with the smallest individual *P* values. No predefined imaging category survived FDR or Bonferroni correction. The smallest category-level FDR-adjusted value was *q* = 0.056 for white-matter-hyperintensity volume. However, this category contained only one phenotype, meaning that its ACAT result was identical to its individual association result.

Resting-state fMRI node amplitude showed suggestive but nonsignificant aggregate evidence:

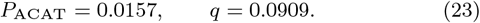

Its median genetic correlation was *r*_*g*_ = *−*0.0507, and 70 of the 75 estimates were negative. The category remained nominally associated after removal of its strongest phenotype, with *P*_ACAT_ = 0.0266.

Regional T2* measures also showed suggestive but nonsignificant aggregate evidence:

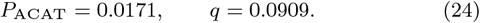

Twelve of the 14 estimates were positive, although the ACAT result weakened to *P*_ACAT_ = 0.0656 after removal of the leading phenotype. These category-level patterns were considered exploratory because their corrected *P* values exceeded 0.05 and because simple direction counts do not account for correlations among imaging phenotypes.

### Power and Precision

The median minimum detectable absolute genetic correlation at 80% power was

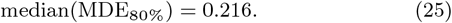

No phenotype had 80% power to detect an absolute genetic correlation of |*r*_*g*_| = 0.10, and only 1.5% had sufficient power to detect |*r*_*g*_| = 0.15. In contrast,

- 38.6% were powered to detect |*r*_*g*_| = 0.20;
- 67.4% were powered to detect |*r*_*g*_| = 0.25; and
- 81.4% were powered to detect |*r*_*g*_| = 0.30.

Bonferroni-adjusted simultaneous confidence intervals were fully contained within

- [*−*0.15, 0.15] for 5.4% of phenotypes;
- [*−*0.20, 0.20] for 38.2%;
- [*−*0.25, 0.25] for 65.4%; and
- [*−*0.30, 0.30] for 80.0%.

Thus, small genetic correlations could not generally be excluded, whereas moderate-to-large correlations were inconsistent with the data for a substantial proportion of imaging phenotypes.

### Sensitivity Analyses

The broad sensitivity analysis included 3,703 biological phenotypes with valid genetic-correlation estimates. No individual phenotype survived correction for multiple comparisons, and the global ACAT test remained nonsignificant:

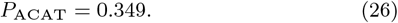

The stringent sensitivity analysis included 2,690 phenotypes with brain SNP-heritability *Z* statistics of at least 3. Again, no individual phenotype or imaging category survived correction, and the global ACAT test was nonsignificant:

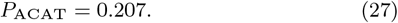

The median genetic correlation remained approximately 0.0014 under the stringent definition. Bonferroni-adjusted simultaneous confidence intervals were contained within [*−*0.30, 0.30] for 88.0% of phenotypes meeting the stringent quality-control criteria.

The stability of the results across the broad, primary, and stringent definitions indicated that the principal conclusion did not depend on the precise SNP-heritability quality-control threshold.

## Discussion

In this comprehensive analysis of nearly 3,000 quality-control-eligible biological UK Biobank brain imaging phenotypes, we found no individual imaging phenotype with a statistically robust genome-wide genetic correlation with externalizing. We also found no significant aggregate evidence across all imaging phenotypes or within predefined structural, diffusion, susceptibility-weighted, resting-state functional, or task functional MRI categories.

The distribution of estimated genetic correlations was centered near zero, and the median absolute genetic correlation was small. These findings suggest that externalizing does not share widespread moderate-to-large directional common-variant effects with individual adult brain imaging phenotypes. This conclusion was consistent across broad, primary, and stringent phenotype-inclusion criteria.

The results should not be interpreted as proof that externalizing and brain phenotypes have no shared genetic influences. The median minimum detectable genetic correlation was approximately 0.22, and the analyses had little power to identify correlations of 0.10–0.15 after correction for almost 3,000 comparisons. Small shared genetic effects may therefore remain undetected.

A genome-wide genetic correlation can also obscure more complex forms of genetic overlap. For example, shared variants with opposing directions of effect across genomic regions may produce a global genetic correlation close to zero. Genetic overlap could also be localized to a small number of genomic regions, distributed across multiple correlated imaging phenotypes, or mediated through developmental or environmental processes.

The age difference between the externalizing GWAS and the UK Biobank imaging sample may also be relevant. Externalizing liability often emerges during childhood, adolescence, or early adulthood, whereas UK Biobank brain imaging was collected primarily from middle-aged and older adults. Genetic effects on brain development may not be represented by the same imaging measures or effect magnitudes later in life.

Although no corrected associations were identified, several exploratory patterns may guide future hypothesis-driven work. Resting-state node-amplitude phenotypes showed predominantly negative estimates and a nominal category-level ACAT result. Regional T2* measures showed predominantly positive estimates. Nominal individual findings also involved thalamic volume, putamen T2*, cerebral-peduncle microstructure, inferior frontal morphology, and resting-state connectivity.

These observations should not be treated as discoveries. Instead, they may be used to formulate hypotheses for independent replication, particularly when supported by previous literature or independent adolescent imaging data. Testing only these regions again in the same dataset would not provide independent confirmation.

The simultaneous-confidence-bound analysis strengthens the interpretation of the null findings. In the primary analysis, genetic correlations larger than 0.30 in absolute magnitude were excluded for 80.0 % of phenotypes. Under the stringent heritability quality-control definition, this proportion increased to 88.0%. Therefore, although small effects remain plausible, widespread large genetic correlations appear unlikely.

## Limitations

Several limitations should be considered. First, imaging GWAS sample sizes were substantially smaller than the externalizing GWAS sample, limiting the precision of many genetic-correlation estimates. Second, the large number of correlated imaging phenotypes imposed a stringent multiple-testing burden. Although FDR correction was less conservative than Bonferroni correction, small true correlations may still have remained undetected. Third, cross-trait LDSC estimates genome-wide directional genetic correlation and may miss local or mixed-direction genetic sharing. Fourth, the externalizing GWAS may have included UK Biobank participants. Although the cross-trait LDSC intercept allows for sample overlap, the exact extent of overlap should be documented. Fifth, ACAT identifies aggregate evidence from P values but does not estimate a common category effect or model effect directions. Finally, the UK Biobank imaging participants are not representative of the full general population and are older than populations in which externalizing behavior commonly develops.

## Conclusion

Across 2,967 quality-control-eligible biological UK Biobank brain imaging phenotypes, no statistically robust individual or category-level genome-wide genetic correlation with externalizing was identified. The results were consistent across alternative quality-control definitions and provide evidence against widespread moderate-to-large global genetic correlations.

However, the study had limited sensitivity to small effects. Small, localized, mixed-direction, or developmentally specific genetic relationships between externalizing and brain phenotypes therefore remain possible. Future studies using larger imaging GWAS samples, developmental neuroimaging cohorts, local genetic-correlation methods, and independent replication will be needed to evaluate these possibilities.

## Data Availability

Code. The analysis code and scripts used in this study are publicly
available at: https://github.com/mw742/ext_ukb_imggwas
Data. This study uses data from the Adolescent Brain
Cognitive Development (ABCD) Study (https://abcdstudy.org),
available through the NIMH Data Archive (NDA). The ABCD data release used was version 5.1. The study is supported by
the National Institutes of Health (NIH) and additional federal
partners under multiple award numbers, including U01DA041048
and U01DA050987. A full list of funders is available at https:
//abcdstudy.org/federal-partners.html.

## Data availability

**Code**. The analysis code and scripts used in this study are publicly available at: https://github.com/mw742/ext_ukb_imggwas

**Data**. This study uses data from the Adolescent Brain Cognitive Development (ABCD) Study (https://abcdstudy.org), available through the NIMH Data Archive (NDA). The ABCD data release used was version 5.1. The study is supported by the National Institutes of Health (NIH) and additional federal partners under multiple award numbers, including U01DA041048 and U01DA050987. A full list of funders is available at https://abcdstudy.org/federal-partners.html.

## Author contributions statement

Mengman Wei conceived the study, designed the analytical framework, performed data processing, statistical analyses, and computational modeling, and drafted the manuscript. Mengman Wei also carried out code implementation, data curation, and interpretation of the results. Qian Peng provided supervision, overall guidance, resource support, and funding acquisition.

## Preprint Notice

This manuscript is a preprint and has not yet undergone peer review. The content is shared to disseminate findings and establish a precedent. Additional analyses and revisions may be incorporated in future versions.

